# COVID-19 pandemic increased the magnitude of mortality risks associated with cold temperature in Italy: A nationwide time-stratified case-crossover study

**DOI:** 10.1101/2020.09.15.20194944

**Authors:** Wenhua Yu, Rongbin Xu, Tingting Ye, Chunlei Han, Shanshan Li, Yuming Guo

## Abstract

**Backgrounds:** The coronavirus disease 2019 (COVID-19) pandemic and some containment measures have changed many people’s lives and behaviours. Whether the pandemic could change the association between cold temperature and mortality remains unknown.

**Objectives:** We aimed to assess whether the association between cold temperature and all-cause mortality in the pandemic period has changed compared to non-COVID-19 period (2015-2019) in Italy.

**Methods:** We collected daily all-cause mortality data and meteorological data for 107 Italian provinces from 1, January 2015 to 31, May 2020. A time-stratified case-crossover design with the distributed lag non-linear model was used to examine the association between cold temperature and all-cause mortality during the first three months (from March to May in 2020) of the COVID-19 outbreak and the same months in 2015-2019.

**Results:** The relative risk (RR) of all-cause mortality at extreme cold temperature (2.5^th^ percentile of temperature at 3 *°C)* in comparison with the minimum mortality temperature (24 °C) was 4.75 [95% confidence interval (CI): 3.90-5.79] in the pandemic period, which is more than triple higher than RR [1.41 (95%CI: 1.33-1.50)] in the same months during 2015-2019. The shift in cold-mortality association was particularly significant for people aged 65-74 years [RR (95%CI): 5.98 (3.78-9.46) in 2020 versus 1.29 (1.10-1.51) in 2015-2019], 75-84 years [5.25 (3.79-7.26) versus 1.40 (1.25-1.56)], and ≥ 85 years [5.03 (3.90-6.51) versus 1.52 (1.39-1.66)], but not significant for those aged 0-64 years [1.95 (1.17-3.24) versus 1.24 (1.05-1.48)].

**Conclusion:** The findings suggest that the COVID-19 pandemic enhanced the risk of cold temperature on mortality in Italy, particularly among the elderly people. Further studies are warranted to understand the exact mechanism when detailed data are available.

## 1. Introduction

With the coronavirus disease 2019 (COVID-19) infection sweeping across the globe at a startling speed (Bedford et al. 2020), restrictive social containment measures such as social distance, travel restriction, and regional emergency lockdown, have been widely implemented to limit viral diffusion and reduce deaths from infection. Italy was one of the countries with the largest number of confirmed cases at the beginning of the COVID-19 outbreak. Since the first local transmission started on the 20^th^ of February, the number of reported cases had followed an exponential trend, peaking with 6,557 new cases on the 21^st^ of March. Simultaneously, restrictive lockdown measures were implemented to the whole country from the 9^th^ of March and were not downgraded until the end of May 2020.

The restricted activities have caused profound changes in people’s lives and behaviours. For example, stay-at-home orders would directly decrease outdoor physical activities (Wang et al. 2020) and change the pattern of social contacts (Prem et al. 2020). It may also indirectly lead to the damage of health through delayed health care for acute emergencies (De Filippo et al. 2020; De Rosa et al. 2020), exacerbations of chronic diseases (Wang and Zhang 2020), and psychological depression (Weinberger et al. 2020). Moreover, the effect of the pandemic on overall population mortality is unknown. Several recent studies (Banerjee et al. 2020; Weinberger et al. 2020) reported that the overall direct and indirect deaths related to COVID-19 may be underestimated in the pandemic period due to the social containment and mitigation activities. Therefore, it is worthwhile to assess the impact of the pandemic on the overall population health in Italy.

The association between ambient temperature and mortality/morbidity outcomes has been well-documented (Gasparrini et al. 2015; Guo et al. 2014). Apart from hypothermia, many adverse health outcomes like cardiovascular and respiratory diseases are associated with both cold and hot temperatures (Guo et al. 2016a; Zhao et al. 2019a; Zhao et al. 2019b). However, the nature of the temperature-mortality relationship in different climates and populations may exhibit significant variation (Carder et al. 2005; Gasparrini et al. 2015). Generally, the non-linear temperature-mortality association is mainly described as a J-, V- or U-shaped curve in different regions and countries worldwide (Gasparrini et al. 2015; Guo et al. 2013; Guo et al. 2014; Ryti et al. 2016). The association could be affected by factors like geographic locations, climatological and socioeconomic conditions, and human behaviours (Gasparrini et al. 2015). Therefore, it is warranted to explore the association between the temperature and the mortality during the pandemic period when people’s lives and behaviours have been hugely altered due to the restrictive social interventions.

Several previous assessments have focused on the effects of temperature variation on the death of COVID-19 (Briz-Redón and Serrano-Aroca 2020; Ma et al. 2020; Xie and Zhu 2020; Yao et al. 2020). However, the number of COVID-19 deaths in official reports is likely underestimated due to improper COVID-19 death certification or the lack of testing availability (Gill and DeJoseph 2020). As a result, many deaths may suffer from misclassification to another respiratory disease, such as influenza. Unlike the deaths attributed to COVID-19, all-cause mortality could be an accurate measurement to estimate the direct and indirect effects of the pandemic on deaths (Zylke and Bauchner 2020). Apart from avoiding the common misclassification of cause-specific mortality, all-cause mortality incorporates both the deaths from the virus and the mortality indirectly from the pandemic. For example, the incidence of out-of-hospital cardiac arrest was significantly higher during the pandemic period in Italy than that at the same time in 2019 (Baldi et al. 2020; Marijon et al. 2020). Such indirect deaths from the pandemic may increase because of the avoidance of health care or the lack of social supports, especially for the vulnerable population who are concerned about being infected when going to a hospital (Baum and Schwartz 2020; Marijon et al. 2020; Rosenbaum 2020). The overlap of vulnerable populations in COVID-19 and temperature-related diseases makes the measurement of the pandemic impacts complexed. Thus, all-cause mortality could be a useful tool to comprehensively assess the impact of temperature on human health in the pandemic.

In this study, we aimed to identify the risk of all-cause mortality associated with temperature in the early spread period of COVID-19 across the 107 Italian provinces between March 1^st^, 2020 and May 31^st^, 2020, and compare the association with that during the same period in 2015-2019. We also performed stratified analyses by sex and age to estimate the influence of temperature on different population in the pandemic period in Italy.

## 2. Methods

### 2.1 Data collection

We collected daily all-cause mortality data for 7,904 municipalities in 107 provinces of Italy from the Italian Institute of Statistics from January 1^st^, 2015 to May 31^st^, 2020, which covered about 93.1% of the Italian territory and population. The mortality counts were grouped into province-level stratified by sex and age. To compare the effects of temperature on mortality before and during the COVID-19 pandemic, the analysis was restricted to the first three months of the COVID-19 outbreak in Italy in 2020 and the same months during 2015-2019, respectively.

We collected the hourly ambient temperature (at 2 meters above the land surface) and ambient dew point temperature at 0.1°x0.1° spatial resolution from the ERA5 dataset (ERA5) (https://cds.climate.copernicus.eu/cdsapp#!/home). We aggregated all hourly observations into daily data. This dataset has good temporal and spatial coverage in Italy, but it might be less accurate than observations from weather stations. Thus, we calibrated the ERA5 daily mean temperature and ERA5 daily mean dew point temperature using their relationship with observations of weather stations by building random forest models (see Supplementary materials for detail, Fig. S1-S2). After calibration, the ERA5 daily mean temperature and ERA5 daily mean dew point temperatures were linked to the geographical centre of each of the 7,904 municipalities based on longitude and latitude. We then calculated daily mean relative humidity from the calibrated ERA5 daily mean temperature and ERA5 daily mean dew point temperature using an algorithm provided by the “humidity” R package (Cai 2019). Weather data at municipality level were aggregated into province level by averaging observations of all municipalities within each province. We also collected the daily monitor stations data of particulate matter with diameters less than 2.5 micrometres (PM_2.5_) from World’s Air Quality Index Project and linked values of the nearest monitor station to the centre of each Italian province.

### 2.2 Statistics analysis

A time-stratified case-crossover design was used to examine the associations between temperature and mortality during the first three months of COVID-19 outbreak and the same pre-outbreak period in 2015-2019, respectively. Conditional quasi-Poisson regression was applied to perform the time-stratified case-crossover design (Armstrong et al. 2014). Time-stratified case-crossover design controls the temporal trends and seasonality through matching case and control in a small time window (e.g., calendar month) (Guo et al. 2011). Specifically, the case and controls were matched by the day of the week in the same calendar month in the same province, to control for the effect of intra-week variation, seasonality, and spatial variation. The association between ambient temperature and risk of death was examined using a distributed lag non-linear model (DLNM) (Gasparrini et al. 2010). We used a natural cubic spline with 3 degrees of freedom for the temperature distribution, and a natural cubic spline with 3 degrees of freedom for lags up to 21 days. We chose a lag of 21 days, as previous studies have shown a long lag effect of cold temperature and harvesting effect of hot temperature (Gasparrini et al. 2015; Guo et al. 2011). The daily relative humidity with the same lag days and PM_2.5_ concentration were included in the model to control their impacts.

To assess the possible modification effects of demographic factors on the associations between temperature and mortality, stratified analyses were conducted for sex and age groups (<64, 65-74, 75-84, and ≥85 years). Our initial analyses show there were only cold effects on mortality (no hot effects). Therefore, the overall cumulative RRs in previous 21 days corresponding to the 2.5^th^ percentile and the 50^th^ percentile of the temperature distribution during the study period against the minimum mortality temperature (MMT) were calculated to show the effects of extreme and moderately cold temperatures, respectively. We set 24°C as MMT (reference value/temperature) based on our initial analyses and a previous study to calculate the relative risks (RRs) (Gasparrini et al. 2015).

### 2.3 Sensitivity analysis

Sensitivity analyses were performed to examine the robustness of the results. We tested the variation of the temperature-mortality association during non-outbreak of COVID- 19 by replacing the study period of 2015-2019 with every single year of the study period. We modified the lag choices by varying the maximum lags to 18 and 24 days in the DLNM to check any changes in the cold temperature-mortality associations.

All analyses were performed with R software (version 3.5.2). The ‘dlnm’ and ‘gnm’ packages were used to fit the DLNM and the conditional Poisson regression, respectively.

## 3. Results

There were 1,000,459 all-cause deaths from March 1^st^ to May 31^st^ during 2015-2020 in Italy. Table 1 shows the comparison of descriptive statistics between the COVID-19 pandemic period in 2020 and the same months during 2015-2019. The study months in the pandemic period in Italy witnessed a slight increase in the ambient temperature (P<0.001) with the average value of 12.5 ± 4.4 *°C* compared with the same months during 2015-2019 (12.3 ± 4.3 °C). In addition, the relative humidity and PM_2.5_ in the pandemic period were significantly lower than the same months during 2015-2019. As for the comparison of the daily average all-cause mortality counts in different age groups, all age groups displayed statistical higher counts in 2020 than the number in 2015-2019, except for the 0-64 age group. The death counts in the pandemic period experienced a significantly high number in male and female groups.

**Table 1.** Descriptive statistics for temperature, relative humidity, PM_2.5_, and daily death counts during COVID-19 pandemic in 2020 and the same months during 2015-2019.

| Characteristics | 2015-2019 |  | 2020 |  | *p*-Value |
| --- | --- | --- | --- | --- | --- |
|  | Mean | SD | Mean | SD |  |
| Temperature (°C) | 12.35 | 4.29 | 12.52 | 4.40 | <0.001 |
| Relative Humidity (%) | 68.80 | 11.15 | 65.41 | 12.08 | <0.001 |
| PM<sub>2.5</sub> (µg/m<sup>3</sup>) | 16.56 | 13.01 | 13.86 | 9.96 | <0.001 |
| Daily Average Mortality counts |  |  |  |  |  |
| 0-64 years | 192 | 17 | 196 | 37 | 0.097 |
| 65-74 years | 235 | 21 | 285 | 84 | <0.001 |
| 75-84 years | 519 | 45 | 634 | 204 | <0.001 |
| ≥ 85 years | 802 | 78 | 1021 | 277 | <0.001 |
| Female | 911 | 80 | 1093 | 271 | <0.001 |
| Male | 837 | 62 | 1043 | 325 | <0.001 |
| Total | 1748 | 135 | 2136 | 590 | <0.001 |

Note: 2020: the pandemic period from March 1st to May 31<sup>st</sup> 2020; 2015-2019: the normal period from March 1st to May 31st for 2015-2019; SD: Standard deviation; PM<sub>2.5</sub>: particulate matter with diameters less than 2.5 micrometres; Daily temperature and relative humidity were calculated with the 24h average values; p-Value was calculated with independent Two-Sample t-Test.

Figure 1 shows overall cumulative exposure-response associations for the pandemic period in 2020 and the same pre-outbreak period in 2015-2019, referring to the MMT of 24°C. Generally, there were only cold effects on mortality for both 2020 COVID-19 pandemic and the same months during 2015-2019. The risks of excess mortality associated with cold temperatures in the pandemic period were significantly higher than that in the 2015-2019 period. A similar trend can be found in sex and age groups (Figure 2). There were similar temperature-mortality curves for males and females. As for the age-specific temperature-mortality association, except for the 0-64 age group, the other three older age groups experienced a significantly higher risk of death in the pandemic period than 2015-2019.

**Fig. 1.**
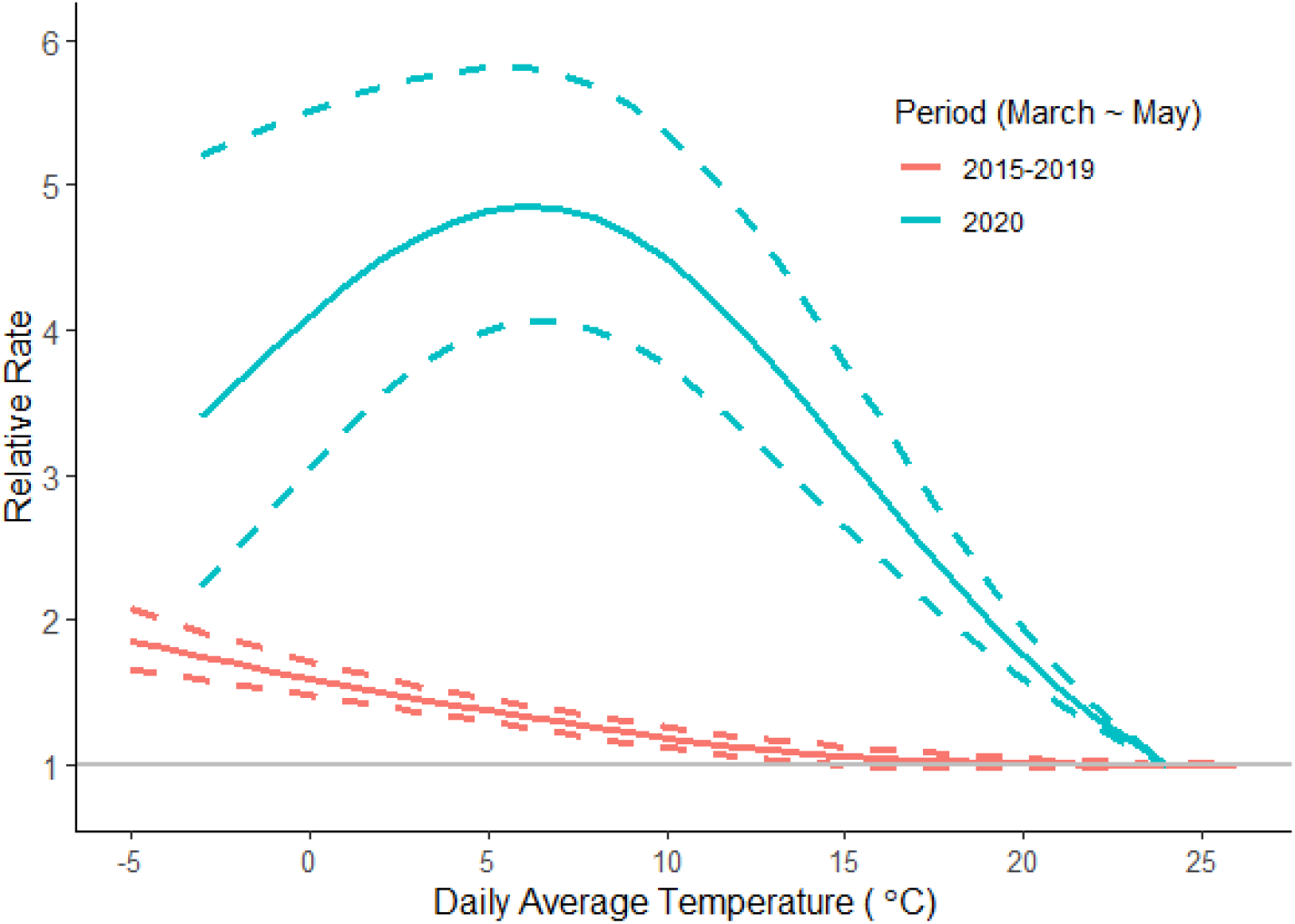
Cumulative temperature-mortality associations along 21 lag days in 2020 COVID-19 pandemic and the same months during 2015-2019.

The cumulative RR of all-cause mortality associated with extreme cold (2.5^th^ percentile of temperature at 3 °C) in comparison with the MMT (24 °C) was 4.75 (95%CI: 3.90-5.79) in the pandemic period, which is more than three times higher than the 2015-2019 period with RR of 1.41 (95%CI: 1.33-1.50). With regards to the association of mortality in moderately cold temperature (50^th^ percentile of temperature at 14°C), Italy had a cumulative RR of 3.75 (95%CI: 3.12-4.52) in the pandemic period compared with that in the non-pandemic period with an RR of 1.10 (95%CI: 1.03-1.17). When compared the risks at extreme and moderately cold temperatures in specific-age groups in the pandemic period, the 65-74 age group reported significantly higher risks than the other three groups at extreme and moderately cold temperatures in the pandemic, while the 0-64 age group showed the similar RR values at these two temperatures (Table 2).

**Table 2.**
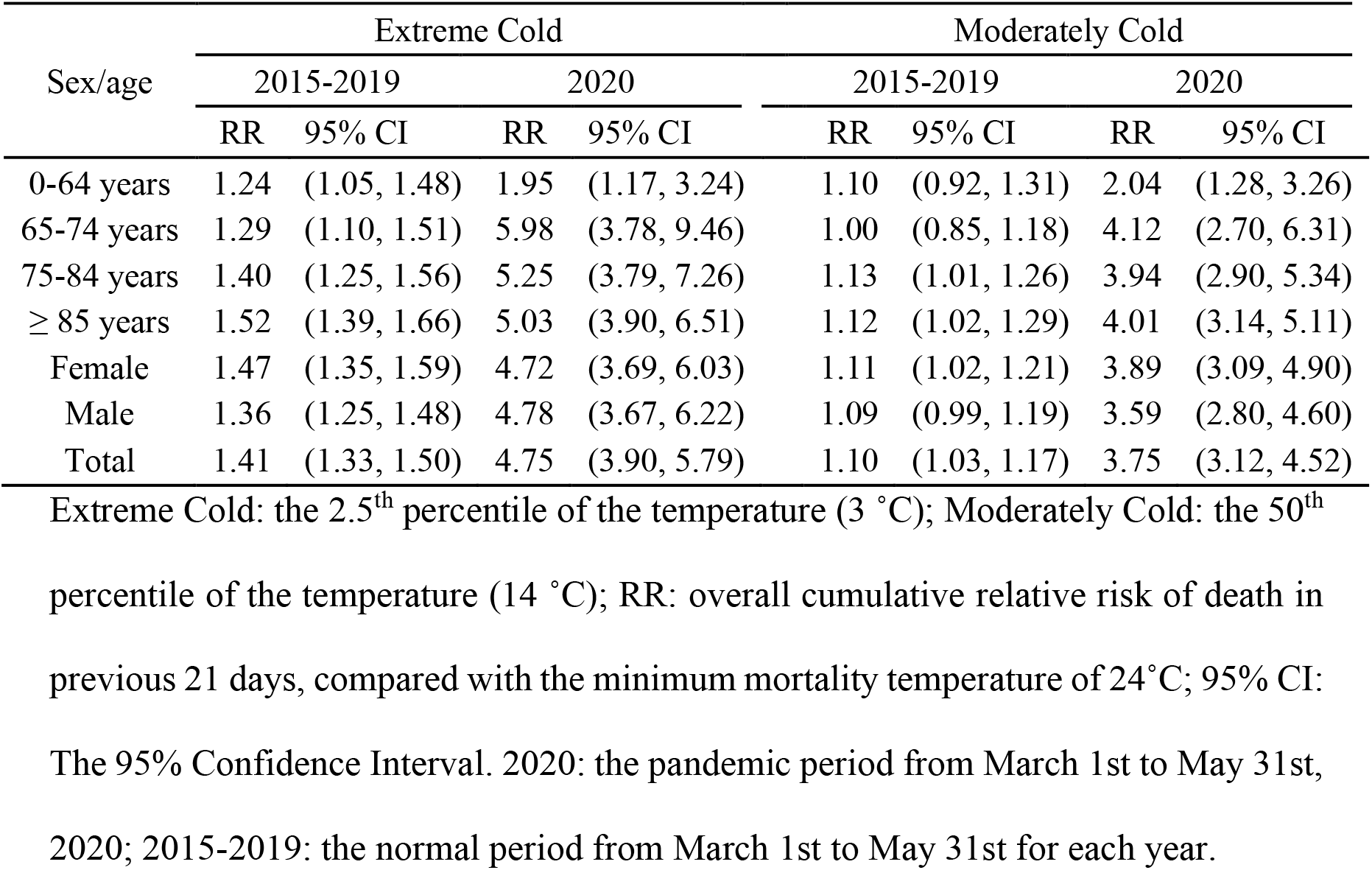
Relationship between temperature and mortality at extreme and moderately cold temperatures.

| Sex/age | Extreme Cold |  |  |  | Moderately Cold |  |  |  |
| --- | --- | --- | --- | --- | --- | --- | --- | --- |
|  | 2015-2019 |  | 2020 |  | 2015-2019 |  | 2020 |  |
|  | RR | 95% CI | RR | 95% CI | RR | 95% CI | RR | 95% CI |
| 0-64 years | 1.24 | (1.05, 1.48) | 1.95 | (1.17, 3.24) | 1.10 | (0.92, 1.31) | 2.04 | (1.28, 3.26) |
| 65-74 years | 1.29 | (1.10, 1.51) | 5.98 | (3.78, 9.46) | 1.00 | (0.85, 1.18) | 4.12 | (2.70, 6.31) |
| 75-84 years | 1.40 | (1.25, 1.56) | 5.25 | (3.79, 7.26) | 1.13 | (1.01, 1.26) | 3.94 | (2.90, 5.34) |
| ≥ 85 years | 1.52 | (1.39, 1.66) | 5.03 | (3.90, 6.51) | 1.12 | (1.02, 1.29) | 4.01 | (3.14, 5.11) |
| Female | 1.47 | (1.35, 1.59) | 4.72 | (3.69, 6.03) | 1.11 | (1.02, 1.21) | 3.89 | (3.09, 4.90) |
| Male | 1.36 | (1.25, 1.48) | 4.78 | (3.67, 6.22) | 1.09 | (0.99, 1.19) | 3.59 | (2.80, 4.60) |
| Total | 1.41 | (1.33, 1.50) | 4.75 | (3.90, 5.79) | 1.10 | (1.03, 1.17) | 3.75 | (3.12, 4.52) |

Extreme Cold: the 2.5<sup>th</sup> percentile of the temperature (3 °C); Moderately Cold: the 50<sup>th</sup> percentile of the temperature (14 °C); RR: overall cumulative relative risk of death in previous 21 days, compared with the minimum mortality temperature of 24°C; 95% CI: The 95% Confidence Interval. 2020: the pandemic period from March 1st to May 31st, 2020; 2015-2019: the normal period from March 1st to May 31st for each year.

**Fig. 2.**
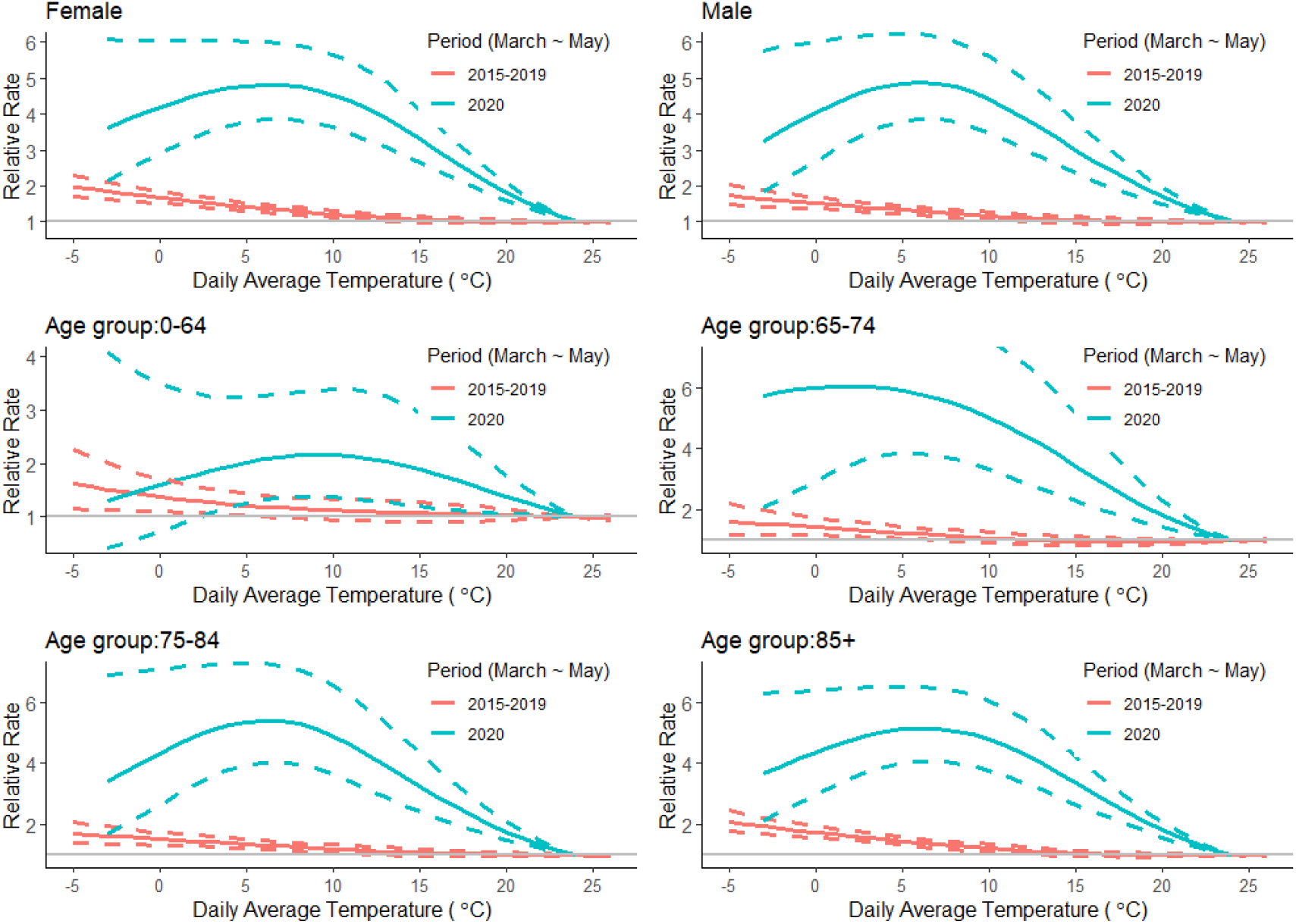
Cumulative exposure-response associations along 21 lag days stratified by sex and age in the pandemic (2020) and the non-pandemic period (2015-2019).

As for the results of sensitivity analysis, the distribution of the temperatures in the same period of 2015-2020 was similar, except for a slightly low temperature in the year 2019 (Table S1). We replaced the study period of 2015-2019 with every single year from 2015 to 2019 to examine the stability of the association between temperature and allcause mortality in the period with no pandemic, which gave similar results (Fig. S3). Additionally, we changed the maximum lags days to 18 and 24 days in our model, and it did not modify the results (Fig. S4). Consequently, we believe that our models adequately captured the main effects of temperature on mortality.

## 4. Discussion

To the best of our knowledge, our study is the first to investigate the cold-related excess mortality during the COVID-19 pandemic period in Italy, even in the world. The findings indicated that Italy experienced a higher cold-related excess mortality during the pandemic in comparison to the equivalent time in 2015-2019. The mortality risks associated with cold temperatures observed during the pandemic were more than triple higher than risks noted in 2015-2019 at both extreme and moderately cold temperatures. Furthermore, risks in individuals with 0-64 year of age displayed a non-significant difference in the pandemic and non-pandemic periods. However, people aged ≥65 years old were more susceptible to the adverse effects of cold temperature than those <65 years old during the pandemic.

It is well-documented that cold temperature is responsible for the excess mortality (Guo et al. 2016b; Schwartz et al. 2015; Vardoulakis et al. 2014). For example, a quantitative systematic review estimated that cold exposure was associated with an elevated risk of cardiovascular and respiratory mortality (Bunker et al. 2016). Our results for the non-pandemic period are consistent with a previous multi-country epidemiological study (Gasparrini et al. 2015), in which it assessed the all-cause mortality risk attributed to ambient temperature in Italian 11 cities from 1987 to 2010 and observed a slightly low cold-related mortality, with an RR of 1.09 in cold temperature. However, the results could not be directly compared due to the different study period.

Various potential biological mechanisms for the effect of cold temperature on the increased mortality risks have been presented, mainly for cardiovascular and respiratory effects (Song et al. 2017). In the case of the association of cold temperature with cardiovascular mortality, cold exposure could increase the angiotensin-II levels in plasma, platelet and red cell counts, blood viscosity, and arterial pressure, which would lead to increased blood pressure and the elevated risks of hypertension and cardiovascular diseases (Analitis et al. 2008; Keatinge et al. 1984). The mechanisms behind the respiratory effects may be associated with the effect of cold temperature on the reduction of respiratory defences against infection and other immunological reactions (Eccles 2002). Meanwhile, the cold temperature could affect the transmission of viruses and enhance the rates of respiratory virus infection (Pica and Bouvier 2014).

Our results show that more than three times higher cold-related excess mortality during 2020 COVID-19 pandemic than the same months during 2015-2019. Even though the potential drivers for such change cannot be directly derived, a multitude of underlying factors are linked to the variation of the cold-mortality association in the pandemic period. Firstly, the contributions of COVID-19 deaths to the cold-related excess mortality need to be taken into account. According to a recent report, in the first 5 months of 2020 (with the 191,228 total deaths in Italy), 31,763 (16.6%) was of subjects with swab positive for COVID-19, but the mortality contribution varied by different prevalence locations ranging from 28% in high prevalence to 3% in low-prevalence provinces (National Statistical Institute (Istat) and the InstituteSuperiore di Sanità (Iss) 2020). It is noted that the role of ambient temperature in the spread of COVID-19, so far, is not well understood. Obviously, the virus could not subside with the warming of temperature in the world. However, the impact of cold air temperature on COVID-19 is more convincing. Several laboratory experiments have suggested lower temperatures are associated with increased survival of the virus (Casanova et al. 2010). Sajadi and colleagues concluded that the early spread of COVID-19 was consistent with the behavior of a seasonal respiratory by examining the global spread of the COVID-19 (Sajadi et al. 2020). Many recent investigations also demonstrated a positive association between cold temperature and COVID-19 risk. For example, an epidemiologic study with 122 Chinese cities estimated a positive association with a 4.86% increase in the daily number of COVID-19 confirmed cases for each 1 °C rise when temperatures below 3 °C (Xie and Zhu 2020).

In addition, another possible explanation for the association change in the pandemic period could be the adaptation of societies to cold temperature (Gasparrini et al. 2017; Schwartz et al. 2015). Specifically, the variation of temperature-mortality association mainly depends on the combination of two pathways: an intrinsic adaptation by a physiological acclimatisation response, and the extrinsic adaptation due to non-climate driven factors such as socioeconomic and demographic conditions and public health services (Achebak et al. 2020; Vicedo-Cabrera et al. 2018). Especially for the early days of the COVID-19 outbreak, extrinsic factors such as the health-care resource availability could play an important role in the change of the vulnerability. The rapid surge of COVID-19 cases has posed a serious threat to the Italian national health system (Remuzzi and Remuzzi 2020). Because of the lack of medical resources (Ji et al. 2020) and reorganization of the health care systems to the enormous increase in acutely COVID-19 patients, some patients died at home while awaiting admission (Emanuel et al. 2020), which might be caused by cold temperatures (Achebak et al. 2019; Yang et al. 2017). For example, there was a significant decrease in the rate of hospital admission for acute coronary syndrome and acute myocardial infarction during the early stage of the COVID-19 outbreak in Italy (De Filippo et al. 2020; De Rosa et al. 2020).

On the other hand, further important consideration is that the adverse psychosocial effects of pandemics (Guessoum et al. 2020) may be related to the individuals’ intrinsic physiological and psychological vulnerability to cold temperature. For example, research revealed that social exclusion could make people feel cold (Zhong and Leonardelli 2008). Several studies also observed that ambient temperatures are associated with the increase in emergency department visits for mental illness, suicides, and self-reported days of poor mental health (Mullins and White 2019; Xue et al. 2019). Restrictive social containment measures (including stay-at-home orders, quarantine, and isolation) may exacerbate the adverse psychological outcomes and mental disorders, resulting in excess cold-related mortality (Pfefferbaum and North 2020).

We also investigated the discrepancy effects by sex and age groups in the pandemic. The difference in temperature-related mortality between sex and age groups may be largely driven by a thermoregulatory pathway. Some studies suggested that men have larger decreases in core body temperature when exposed to cold, and thus are more susceptible to cold than women, whereas other studies reported the opposite (Achebak et al. 2019; Moghadamnia et al. 2017). The ability to maintain body temperature for males and females may differ with health and socioeconomic conditions (Moghadamnia et al. 2017). With regards to the age-specific differences in thermoregulatory during exposure to cold temperature, older people generally respond with reduced peripheral vasoconstriction and decreased metabolic heat production in comparison with younger individuals (Achebak et al. 2019; Stocks et al. 2004). Additionally, multiple factors other than physiological mechanisms could explain the discrepancies in pandemic cold-related mortality between different age groups. One possible explanation for the pronounced gap in the RRs between the 65-74 age group and other older groups is the health-related behaviours. For example, the older individuals may be more concerned about their vulnerability of COVID-19 infection and are more willing to conduct personal protection measures such as wearing masks and staying indoors and thus avoid direct to cold exposures and infectious diseases.

There are some limitations in this study. First, we did not include the analysis of the disease-specific mortality because the data was not available. Therefore, we did not know the extent of the temperature impact on certain disease-specific mortality in the pandemic. An additional limitation is that this is an ecological study without individual data. Therefore, exposure misclassification is a potential concern. For example, the aggregated ambient temperature at a province-level does not necessarily remain consistent for individuals, specifically for people who may reduce the cold exposure because of keeping the stay-at-home orders during the pandemic. Furthermore, we were unable to make any inference in terms of the possible mechanism pathways governing the association between cold exposure and excess mortality in the pandemic.

In conclusion, our findings suggest that COVID-19 pandemic increased the impacts of cold temperatures on mortality in Italy. This means that using the historical exposure-response relationship between temperature and mortality may underestimate the health impacts of cold temperatures during the COVID-19 pandemic. To date, no sign shows the end of the COVID-19 pandemic before the widespread use of vaccines. Some restrictive social containment measures may continually affect our life and behaviour in the foreseeable future. Therefore, we would expect our findings enable public health agencies to take early actions such as making preparedness for indoor temperature control, developing early cold warning systems, and giving special attention to the vulnerable group, to avoid the unexpected cold-related excess mortality in the next coming winter.

## Data Availability

We collected daily all-cause mortality data for 7,904 municipalities in 107 provinces of Italy from the Italian Institute of Statistics; We collected the hourly ambient temperature and ambient dew point temperature from the ERA5 dataset (ERA5) (https://cds.climate.copernicus.eu/cdsapp#!/home).

https://cds.climate.copernicus.eu/cdsapp#!/hom

https://www.istat.it

## Reference

Achebak H, Devolder D, Ballester J. 2019. Trends in temperature-related age-specific and sex-specific mortality from cardiovascular diseases in spain: A national time-series analysis. The Lancet Planetary Health 3:e297-e306.

Achebak H, Devolder D, Ingole V, Ballester J. 2020. Reversal of the seasonality of temperature-attributable mortality from respiratory diseases in spain. Nature communications 11:1–9.

Analitis A, Katsouyanni K, Biggeri A, Baccini M, Forsberg B, Bisanti L, et al. 2008. Effects of cold weather on mortality: Results from 15 european cities within the phewe project. American journal of epidemiology 168:1397–1408.

Armstrong BG, Gasparrini A, Tobias A. 2014. Conditional poisson models: A flexible alternative to conditional logistic case cross-over analysis. BMC medical research methodology 14:122.

Baldi E, Sechi GM, Mare C, Canevari F, Brancaglione A, Primi R, et al. 2020. Out-of-hospital cardiac arrest during the covid-19 outbreak in italy. New England Journal of Medicine.

Banerjee A, Pasea L, Harris S, Gonzalez-Izquierdo A, Torralbo A, Shallcross L, et al. 2020. Estimating excess 1-year mortality associated with the covid-19 pandemic according to underlying conditions and age: A population-based cohort study. The Lancet.

Baum A, Schwartz MD. 2020. Admissions to veterans affairs hospitals for emergency conditions during the covid-19 pandemic. JAMA.

Bedford J, Enria D, Giesecke J, Heymann DL, Ihekweazu C, Kobinger G, et al. 2020. Covid- 19: Towards controlling of a pandemic. The Lancet 395:1015–1018.

Briz-Redón Á, Serrano-Aroca Á. 2020. A spatio-temporal analysis for exploring the effect of temperature on covid-19 early evolution in spain. Science of the Total Environment:138811.

Bunker A, Wildenhain J, Vandenbergh A, Henschke N, Rocklöv J, Hajat S, et al. 2016. Effects of air temperature on climate-sensitive mortality and morbidity outcomes in the elderly; a systematic review and meta-analysis of epidemiological evidence. EBioMedicine 6:258–268.

Cai J. 2019. Humidity: Calculate water vapor measures from temperature and dew point. Available: https://cran.r-project.org/web/packages/humidity/index.html.

Carder M, McNamee R, Beverland I, Elton R, Cohen G, Boyd J, et al. 2005. The lagged effect of cold temperature and wind chill on cardiorespiratory mortality in scotland. Occupational and environmental medicine 62:702–710.

Casanova LM, Jeon S, Rutala WA, Weber DJ, Sobsey MD. 2010. Effects of air temperature and relative humidity on coronavirus survival on surfaces. Applied and environmental microbiology 76:2712–2717.

De Filippo O, D’Ascenzo F, Angelini F, Bocchino PP, Conrotto F, Saglietto A, et al. 2020. Reduced rate of hospital admissions for acs during covid-19 outbreak in northern italy. New England Journal of Medicine.

De Rosa S, Spaccarotella C, Basso C, Calabrò MP, Curcio A, Filardi PP, et al. 2020. Reduction of hospitalizations for myocardial infarction in italy in the covid-19 era. European heart journal 41:2083–2088.

Eccles R. 2002. An explanation for the seasonality of acute upper respiratory tract viral infections. Acta oto-laryngologica 122:183–191.

Emanuel EJ, Persad G, Upshur R, Thome B, Parker M, Glickman A, et al. 2020. Fair allocation of scarce medical resources in the time of covid-19. Mass Medical Soc.

Gasparrini A, Armstrong B, Kenward MG. 2010. Distributed lag non - linear models. Statistics in medicine 29:2224–2234.

Gasparrini A, Guo Y, Hashizume M, Lavigne E, Zanobetti A, Schwartz J, et al. 2015. Mortality risk attributable to high and low ambient temperature: A multicountry observational study. The Lancet 386:369–375.

Gasparrini A, Guo Y, Sera F, Vicedo-Cabrera AM, Huber V, Tong S, et al. 2017. Projections of temperature-related excess mortality under climate change scenarios. The Lancet Planetary Health 1:e360-e367.

Gill JR, DeJoseph ME. 2020. The importance of proper death certification during the covid-19 pandemic. JAMA.

Guessoum SB, Lachal J, Radjack R, Carretier E, Minassian S, Benoit L, et al. 2020. Adolescent psychiatric disorders during the covid-19 pandemic and lockdown. Psychiatry research:113264.

Guo Y, Barnett AG, Pan X, Yu W, Tong S. 2011. The impact of temperature on mortality in tianjin, china: A case-crossover design with a distributed lag nonlinear model. Environmental health perspectives 119:1719–1725.

Guo Y, Li S, Zhang Y, Armstrong B, Jaakkola JJ, Tong S, et al. 2013. Extremely cold and hot temperatures increase the risk of ischaemic heart disease mortality: Epidemiological evidence from china. Heart 99:195–203.

Guo Y, Gasparrini A, Armstrong B, Li S, Tawatsupa B, Tobias A, et al. 2014. Global variation in the effects of ambient temperature on mortality: A systematic evaluation. Epidemiology (Cambridge, Mass) 25:781.

Guo Y, Gasparrini A, Armstrong BG, Tawatsupa B, Tobias A, Lavigne E, et al. 2016a. Temperature variability and mortality: A multi-country study. Environmental health perspectives 124:1554–1559.

Guo Y, Li S, Li Liu D, Chen D, Williams G, Tong S. 2016b. Projecting future temperature- related mortality in three largest australian cities. Environmental pollution 208:66–73.

Ji Y, Ma Z, Peppelenbosch MP, Pan Q. 2020. Potential association between covid-19 mortality and health-care resource availability. The Lancet Global Health 8:e480.

Keatinge W, Coleshaw S, Cotter F, Mattock M, Murphy M, Chelliah R. 1984. Increases in platelet and red cell counts, blood viscosity, and arterial pressure during mild surface cooling: Factors in mortality from coronary and cerebral thrombosis in winter. Br Med J (Clin Res Ed) 289:1405–1408.

Ma Y, Zhao Y, Liu J, He X, Wang B, Fu S, et al. 2020. Effects of temperature variation and humidity on the death of covid-19 in wuhan, china. Science of The Total Environment:138226.

Marijon E, Karam N, Jost D, Perrot D, Frattini B, Derkenne C, et al. 2020. Out-of-hospital cardiac arrest during the covid-19 pandemic in paris, france: A population-based, observational study. The Lancet Public Health.

Moghadamnia MT, Ardalan A, Mesdaghinia A, Keshtkar A, Naddafi K, Yekaninejad MS. 2017. Ambient temperature and cardiovascular mortality: A systematic review and meta-analysis. PeerJ 5:e3574.

Mullins JT, White C. 2019. Temperature and mental health: Evidence from the spectrum of mental health outcomes. Journal of health economics 68:102240.

National Statistical Institute (Istat) and the InstituteSuperiore di Sanità (Iss). 2020. Impact of the covid-19 epidemic on total mortality of resident population period january-may 2020. Available: https://www.istat.it/it/archivio/245573.

Pfefferbaum B, North CS. 2020. Mental health and the covid-19 pandemic. New England Journal of Medicine.

Pica N, Bouvier NM. 2014. Ambient temperature and respiratory virus infection. The Pediatric infectious disease journal 33:311–313.

Prem K, Liu Y, Russell TW, Kucharski AJ, Eggo RM, Davies N, et al. 2020. The effect of control strategies to reduce social mixing on outcomes of the covid-19 epidemic in wuhan, china: A modelling study. The Lancet Public Health.

Remuzzi A, Remuzzi G. 2020. Covid-19 and italy: What next? The Lancet.

Rosenbaum L. 2020. The untold toll—the pandemic’s effects on patients without covid- 19. New England Journal of Medicine:Mass Medical Soc.

Ryti NR, Guo Y, Jaakkola JJ. 2016. Global association of cold spells and adverse health effects: A systematic review and meta-analysis. Environmental health perspectives 124:12–22.

Sajadi MM, Habibzadeh P, Vintzileos A, Shokouhi S, Miralles-Wilhelm F, Amoroso A. 2020. Temperature, humidity, and latitude analysis to estimate potential spread and seasonality of coronavirus disease 2019 (covid-19). JAMA Network Open 3:e2011834-e2011834.

Schwartz JD, Lee M, Kinney PL, Yang S, Mills D, Sarofim MC, et al. 2015. Projections of temperature-attributable premature deaths in 209 us cities using a cluster-based poisson approach. Environmental Health 14:85.

Song X, Wang S, Hu Y, Yue M, Zhang T, Liu Y, et al. 2017. Impact of ambient temperature on morbidity and mortality: An overview of reviews. Science of the Total Environment 586:241–254.

Stocks JM, Taylor NA, Tipton MJ, Greenleaf JE. 2004. Human physiological responses to cold exposure. Aviation, space, and environmental medicine 75:444–457.

Vardoulakis S, Dear K, Hajat S, Heaviside C, Eggen B, McMichael AJ. 2014. Comparative assessment of the effects of climate change on heat-and cold-related mortality in the united kingdom and australia. Environmental health perspectives 122:1285–1292.

Vicedo-Cabrera AM, Sera F, Guo Y, Chung Y, Arbuthnott K, Tong S, et al. 2018. A multicountry analysis on potential adaptive mechanisms to cold and heat in a changing climate. Environment international 111:239–246.

Wang H, Zhang L. 2020. Risk of covid-19 for patients with cancer. The Lancet Oncology 21:e181.

Wang X, Lei SM, Le S, Yang Y, Zhang B, Yao W, et al. 2020. Bidirectional influence of the covid-19 pandemic lockdowns on health behaviors and quality of life among chinese adults. International Journal of Environmental Research and Public Health 17:5575.

Weinberger DM, Chen J, Cohen T, Crawford FW, Mostashari F, Olson D, et al. 2020. Estimation of excess deaths associated with the covid-19 pandemic in the united states, march to may 2020. JAMA Internal Medicine.

Xie J, Zhu Y. 2020. Association between ambient temperature and covid-19 infection in 122 cities from china. Science of The Total Environment 724:138201.

Xue T, Zhu T, Zheng Y, Zhang Q. 2019. Declines in mental health associated with air pollution and temperature variability in china. Nature communications 10:1–8.

Yang J, Zhou M, Ou C-Q, Yin P, Li M, Tong S, et al. 2017. Seasonal variations of temperature-related mortality burden from cardiovascular disease and myocardial infarction in china. Environmental Pollution 224:400–406.

Yao Y, Pan J, Liu Z, Meng X, Wang W, Kan H, et al. 2020. No association of covid-19 transmission with temperature or uv radiation in chinese cities. European Respiratory Journal 55.

Zhao Q, Li S, Coelho MS, Saldiva PH, Hu K, Arblaster JM, et al. 2019a. Geographic, demographic, and temporal variations in the association between heat exposure and hospitalization in brazil: A nationwide study between 2000 and 2015. Environmental health perspectives 127:017001.

Zhao Q, Li S, Coelho MS, Saldiva PH, Hu K, Huxley RR, et al. 2019b. The association between heatwaves and risk of hospitalization in brazil: A nationwide time series study between 2000 and 2015. PLoS medicine 16:e1002753.

Zhong C-B, Leonardelli GJ. 2008. Cold and lonely: Does social exclusion literally feel cold? Psychological Science 19:838–842.

Zylke JW, Bauchner H. 2020. Mortality and morbidity: The measure of a pandemic. JAMA.

